# Diagnostic Value of Carotid Ring Sign in Predicting Internal Carotid True Occlusion

**DOI:** 10.1101/2023.09.13.23295519

**Authors:** Tingyu Yi, Yi Sui, Ding-huan Zheng, Xinwen Ren, Xiao-hui Lin, Yan-min Wu, Ding-lai Lin, Zhi-nan Pan, Xiu-fen Zheng, Ganji Hong, Mei-hua Wu, Lisan Zeng, Wen-huo Chen

## Abstract

**BACKGROUND AND PURPOSE:** To differentiate between pseudo-occlusion (PO) and true occlusion (TO) of internal carotid artery (ICA) is important in thrombectomy treatment planning for acute ischemic stroke (AIS) patients. Although delayed contrast filling has been differentiated carotid PO from TO, its application has been limited by the implementations of multiphasic computed tomography angiography (mCTA). In this study, we hypothesized that carotid ring sign, which is readily acquired from single-phasic CTA, can sufficiently differentiate carotid TO from PO.

**METHODS:** Two hundred patients with non-visualization of the proximal ICA were included. Diagnosis of PO or TO of the cervical segment of ICA was made based on digital subtraction angiography. Diagnostic performances of carotid ring sign on arterial-phasic CTA and delayed contrast filling on mCTA were evaluated and compared.

**RESULTS:** One-hundred-twelve patients had ICA PO and 88 had TO. Carotid ring sign was more common in TO patients (70.5% vs 6.3%, p<0.001), while delayed contrast filling was more common in PO (94.9% vs 7.7%, p<0.001). The sensitivity and specificity of carotid ring sign in diagnosing carotid TO was 0.70 and 0.94 respectively, while sensitivity and specificity of delayed contrast filling was 0.95 and 0.92 in judging carotid PO.

**CONCLUSIONS:** Carotid ring sign is a potent imaging marker in diagnosing ICA TO. Carotid ring sign could be complementary to delayed contrast filling sign in differentiating TO from PO, in particular in centers with only single-phasic CTA.

## Introduction

Identification and differentiation of true occlusion (TO) and pseudo-occlusion (PO) of internal carotid artery (ICA) are critical for endovascular management of acute ischemic stroke(AIS)^1,2^. Computed Tomography Angiography (CTA) has been suggested as a reliable tool in assessing thrombus length, clot burden, collaterals and decision making regarding endovascular therapies (EVT)^3–7^. Delayed contrast filling derived from multiphase CTA(mCTA) showed an adequate diagnostic value in differentiating PO from TO^8^. However, the small-scale study may need further verification as visual assessment of faint change of intraluminal contrast staining might be even missed by experienced neurologists^9^. More importantly, multiphasic CTA is not routinely available in many stroke centers. In this study, we hypothesized carotid ring sign derived on single-phasic CTA (sCTA) is a potent imaging marker to differentiate carotid TO from PO.

## Methods

### Patient population and clinical data collection

The study is based on a prospective, hospital-based registry of consecutive large vessel occlusion (LVO) patients treated with endovascular therapies (EVT). Information about patient characteristics, medical and endovascular interventions, periprocedual management and clinical prognosis were routinely collected through an electronic database after patients were admitted to the hospital.

### Definition of TO, PO, carotid ring sign and delayed contrast filling

Digital subtraction angiography (DSA) was used as a reference standard imaging for diagnosis of TO or PO of the proximal cervical ICA. Proximal C1 ICA occlusion on DSA was defined as an actually angiographical occlusion which impedes the passage of contrast to distal C1 segment and the advance of guidewire or catheter^1^, which was also named as ICA TO. Proximal C1 ICA occlusion on CTA was defined as complete nonattenuation of proximal segment of cervical ICA on arterial phase CTA. ICA PO was defined as the demonstration of occlusion on CTA while revealed patent on DSA of proximal C1 ICA. We defined the CTA carotid ring sign as a ring-like appearance > 50% circumference of the vessel wall at the levels of ICA C1 maximum stenosis on axial maximum intensity projection images^10^. The CTA carotid ring sign may be contrast-enhanced or -nonenhanced. We defined the phenomenon of delayed contrast filling as a delayed demonstration of arterial contrast at distal ICA C1 segment during the second or third phase instead of the first phase of mCTA^8^.

### Inclusion and exclusion criteria

Patients were eligible for the study if they met all following criteria: 1) AIS due to LVO in the anterior circulation, 2) demonstration of C1 occlusion on arterial phase CTA, 3) underwent endovascular therapy in acute phase. Patients were excluded from the study if 1) the CTA imaging did not have adequate quality for analysis, 2) the demonstration of C1 occlusion was not the culprit vessel.

### CTA protocol and imaging acquisition

Two computed tomography (CT) machines were used for the study. For Siemens Force scanner, the CTA parameters were: slice thickness 0.75 mm, increment 0.5 mm, tube voltage 80 kV (ref mAs: 129mAs∼72mAs), reconstruction field of view 200 mm. For GE Revolution scanner, the CTA parameters were: slice thickness 0.625 mm, increment 0.625 mm, tube voltage 80∼120 kV, reconstruction field of view 250 mm. For both scanners, automatic bolus tracking technique were applied after the intravenous injection of the contrast (100Hu, flow rate 4.5∼6ml/s, contrast volume: 45∼60ml, NaCl volume: 45∼60ml). For the arterial phase, a contrast agent tracking-triggered approach was utilized. The monitoring site was the ascending aorta at the aortic arch level. After contrast agent injection, there was an 8-second delay before monitoring commenced. Monitoring occurred every 1.5 seconds, with a trigger threshold set at 100 HU(normally 1-3 circles). Once triggered, there was an additional 3-second delay before initiating a cranial-directed arterial phase scan. The scanning range spanned from the tracheal bifurcation to the apex of the skull, with an approximate scanning time of 3 seconds. For the venous phase, there was a 5.5-second delay, with the scanning range extending from the bifurcation of the common carotid artery to the skull apex. For the delay phase, there was a 7-second delay, and the scanning covered from the C1 vertebra to the skull apex.

### Statistical analysis

Quantitative variables were expressed as the mean and SD or median and interquartile range (IQR), and categorical variables were expressed as N number and frequencies (%). The Student t test, Pearson χ ² test, and Cochran Mantel Haenszel χ ² were used to compare the differences between the true and pseudo-occlusion groups. Variables with p values < 0.1 from univariate analysis were adjusted in multivariate analysis model for outcome assessment.

To assess the diagnostic capacity of distinct imaging features for the differentiation between true from pseudo-occlusion, the area under the receiver operating characteristics (ROC) curves were calculated. DeLong approach was used to compare the area under the curves between carotid ring sign, calcification at occlusion sites, and delayed contrast filling. All statistical inferences were two-sided and *p* < .05 was defined as significant. SPSS (version 15.0; SPSS Inc., Chicago, IL, USA) were used for all statistical calculations.

## Results

### Patient characteristics

Patients were retrospectively screened and selected through a prospectively designed hospital-based electronic database (Nalong 2.0, Xiamen, Fujian, China). From June 2018 to February 2022, electrical medical records of 1420 patients with acute LVO in the anterior circulation underwent emergent EVT were consecutively screened (fig 2). Three hundred seventy patients were found to have ICA occlusion on admission CTA, while 211 patients with C1 segment occlusion were included and 159 patients with C2-C7 occlusion were excluded. After exclusion of 7 patients with poor imaging quality and 4 patients with inconclusive neurointerventional procedures. Finally, there were 200 patients in total included in final analysis, among which 69 patients were analyzed for delayed contrast filling using mCTA. Images of single-phase CTA or arterial phase of mCTA from all 200 patients were analyzed for carotid ring sign and calcification at occlusion sites (Fig 1). There were TO in 88 patients and PO in 112 patients.

**Figure 1.**
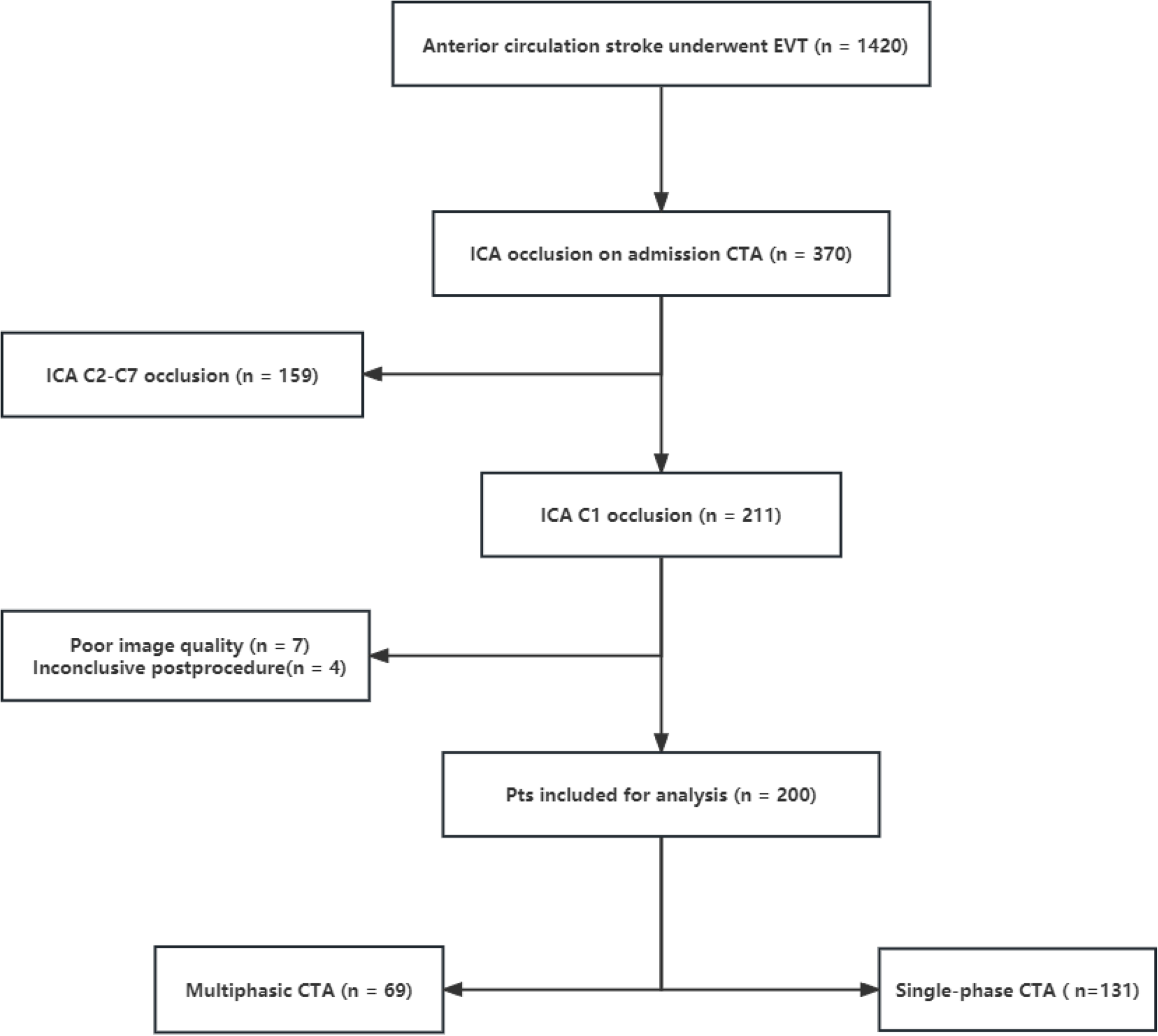
Flowchart of patient selection.

**Figure 2.**
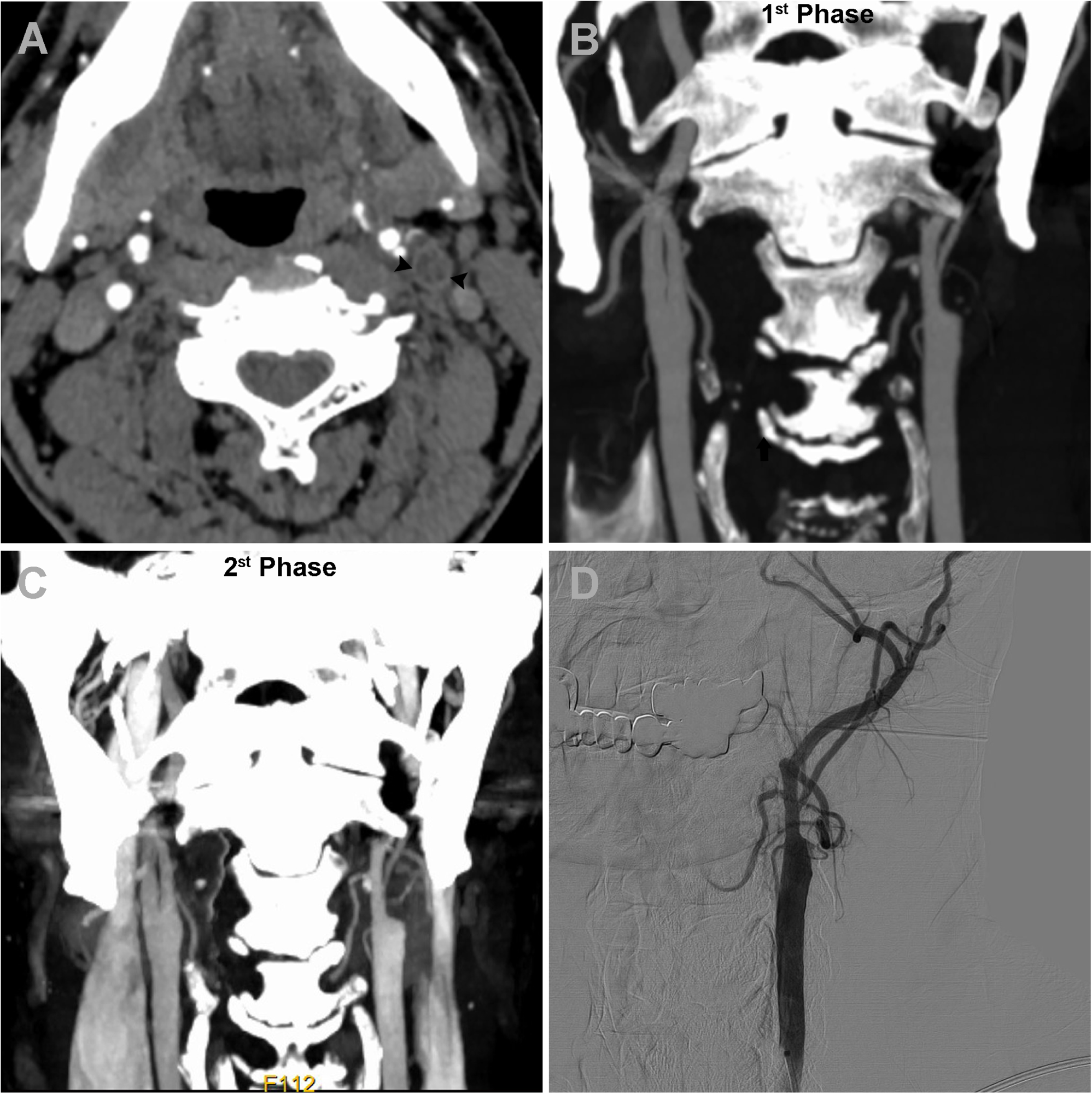
A representative case showing a patient with true occlusion at left ICA C1 segment with positive carotid ring sign and negative delayed contrast filling. A positive carotid ring sign was detected on axial maximum intensity projection image (arrow heads, A). Compared with the 1^st^ phase in mCTA (B), there was no delayed contrast filling revealed in 2^nd^ phase (C).The true left C1 occlusion demonstrated an identical platform-like stump in both CTA phases (C and D), which was confirmed by DSA and endovascular maneuver (D).

### Baseline comparison between TO and PO patients

The main characteristics of patients are summarized in Table 1. Patients with TO were more of male sex compared with PO (77.8 % vs 55.5 %, *p* = 0.001). In line with the finding of more hyperlipidemia in TO patients (26.1% vs 8.9%, *p* < 0.001), atrial fibrillation rate was higher in PO patients (57.1 % vs 8.0 %, *p* < 0.001), etiology of large artery atherosclerosis (LAA) was more frequent in TO patients (92.0 % vs 24.1 %, *p* < 0.001) and cardio-embolism rate was more frequent in PO group (67.9 % vs 2.3 %, *p* < 0.001). Not surprisingly, compared with TO patients, carotid stenting was more frequently deployed in TO patients (92.0% vs 0 %, *p* < 0.001). Three imaging markers showed diagnostic potentials for differentiating PO from TO of ICA C1 segment. Both the presence of carotid ring sign and calcification at the occlusion site were significantly higher in TO compared with PO patients (70.5 % vs 6.3 %, *p* < 0.001; 75.0 % vs 36.6 %, *p* < 0.001). In PO group (n=112), there were only 7 cases with carotid ring sign, i.e., 85.75% (6/7) with hypertension, 85.75% (6/7) with atrial fibrillation, 42.9% (3/7) with diabetes mellitus. Compared with cases without carotid ring sign (n=105), patients with carotid ring sign (n=7) was elder (84 vs 70, *p* < 0.001) and less male sex (0% vs 57.1%, p=0.011).

**Table 1.**
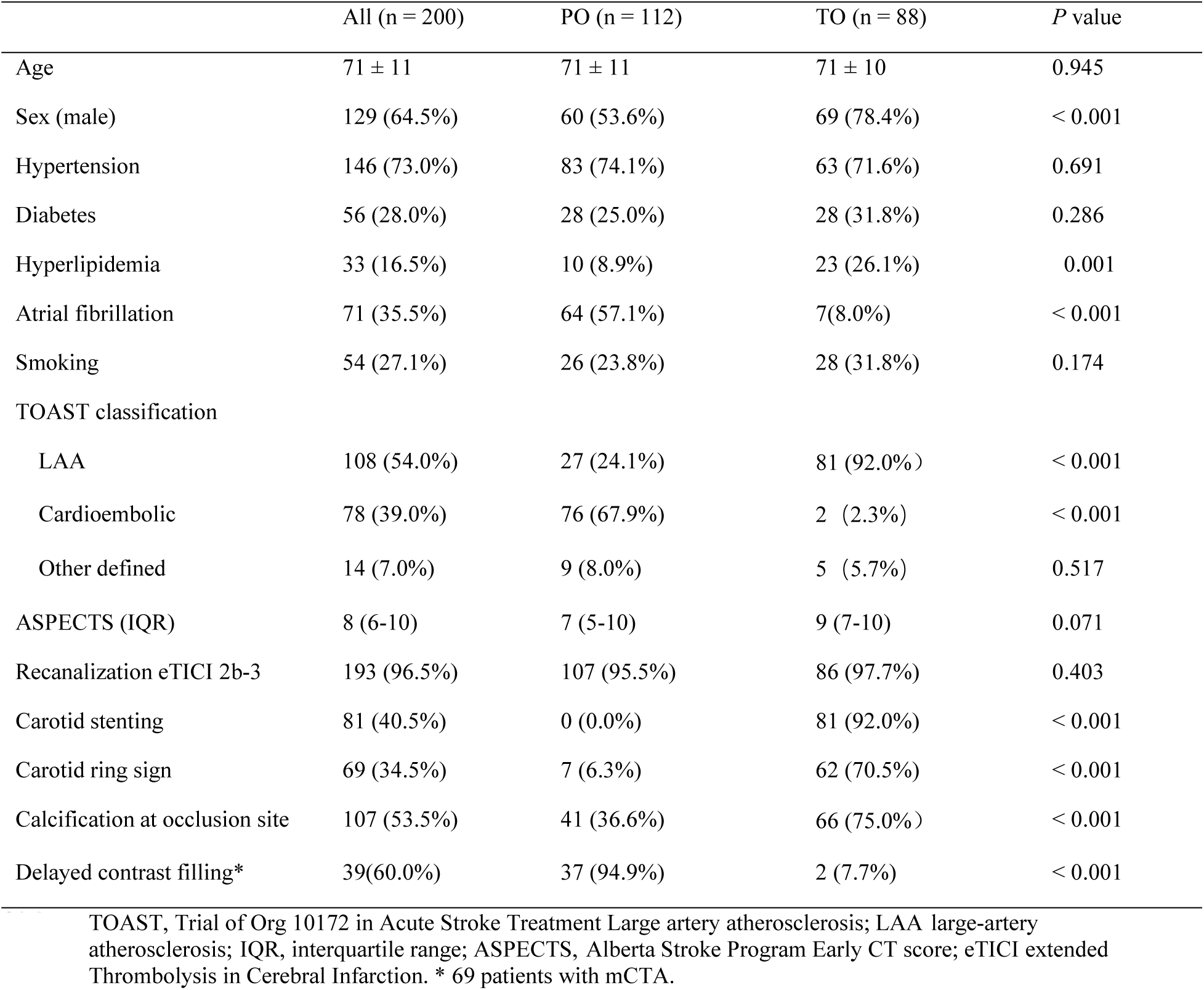
Demographics and characteristics of included patients stratified by pseudo- and true occlusion.

|  | All (n = 200) | PO (n = 112) | TO (n = 88) | P value |
| --- | --- | --- | --- | --- |
| Age | 71 ± 11 | 71 ± 11 | 71 ± 10 | 0.945 |
| Sex (male) | 129 (64.5%) | 60 (53.6%) | 69 (78.4%) | < 0.001 |
| Hypertension | 146 (73.0%) | 83 (74.1%) | 63 (71.6%) | 0.691 |
| Diabetes | 56 (28.0%) | 28 (25.0%) | 28 (31.8%) | 0.286 |
| Hyperlipidemia | 33 (16.5%) | 10 (8.9%) | 23 (26.1%) | 0.001 |
| Atrial fibrillation | 71 (35.5%) | 64 (57.1%) | 7(8.0%) | < 0.001 |
| Smoking | 54 (27.1%) | 26 (23.8%) | 28 (31.8%) | 0.174 |
| TOAST classification |  |  |  |  |
| LAA | 108 (54.0%) | 27 (24.1%) | 81 (92.0%) | < 0.001 |
| Cardioembolic | 78 (39.0%) | 76 (67.9%) | 2 (2.3%) | < 0.001 |
| Other defined | 14 (7.0%) | 9 (8.0%) | 5 (5.7%) | 0.517 |
| ASPECTS (IQR) | 8 (6-10) | 7 (5-10) | 9 (7-10) | 0.071 |
| Recanalization eTICI 2b-3 | 193 (96.5%) | 107 (95.5%) | 86 (97.7%) | 0.403 |
| Carotid stenting | 81 (40.5%) | 0 (0.0%) | 81 (92.0%) | < 0.001 |
| Carotid ring sign | 69 (34.5%) | 7 (6.3%) | 62 (70.5%) | < 0.001 |
| Calcification at occlusion site | 107 (53.5%) | 41 (36.6%) | 66 (75.0%) | < 0.001 |
| Delayed contrast filling* | 39(60.0%) | 37 (94.9%) | 2 (7.7%) | < 0.001 |

TOAST, Trial of Org 10172 in Acute Stroke Treatment Large artery atherosclerosis; LAA large-artery atherosclerosis; IQR, interquartile range; ASPECTS, Alberta Stroke Program Early CT score; eTICI extended Thrombolysis in Cerebral Infarction. \* 69 patients with mCTA.

Totally 69 patients underwent mCTA, and the presence of delayed contrast filling was significantly frequent in PO patients (94.9 % vs 7.7 %, *p* < 0.001). In TO groups (n=88), there were only 2 cases with delayed contrast filling, of which occlusion site located at the middle segment of C1, and very little contrast through C1 on DSA in one case.

### Inter-observer agreement

All imaging features assessed in the study were independently reviewed by 2 experienced neurointerventionists (DH.Z. and ZN.P.) who were blind to results of the interventional angiography. Cases with disagreement were reviewed again and the dispute were resolved by consensus. Cohen kappa coefficient was used to measure inter-observer agreement of various imaging markers. Excellent agreement was achieved (*kappa* =0.90) for both carotid ring sign and delayed contrast filling sign.

### Comparison of diagnostic performance between CTA carotid ring sign and delayed contrast filling

Table 2 summarizes the differential diagnostic values of carotid ring sign for TO and delayed contrast filling for PO. The carotid ring sign has a AUC of 0.86 (SE ± 0.05), sensitivity of 0.70 (95% CI, 0.60-0.79), specificity of 0.94 (95% CI, 0.87-0.97), PPV of 0.90 (95% CI, 0.80-0.95) and NPV of 0.80 (95% CI, 0.72-0.86). The delayed contrast filling sign has an AUC of 0.94 (SE ± 0.04), sensitivity of 0.95 (95% CI, 0.92-0.99), specificity of 0.92 (95% CI, 0.73-0.99), PPV of 0.95 (95% CI, 0.81-0.99) and NPV of 0.92 (95% CI, 0.73-0.99). Representative images of carotid ring sign suggesting TO at C1 segment and delayed contrast filling suggesting PO were presented in Figure 2 and 3 respectively.

**Figure 3.**
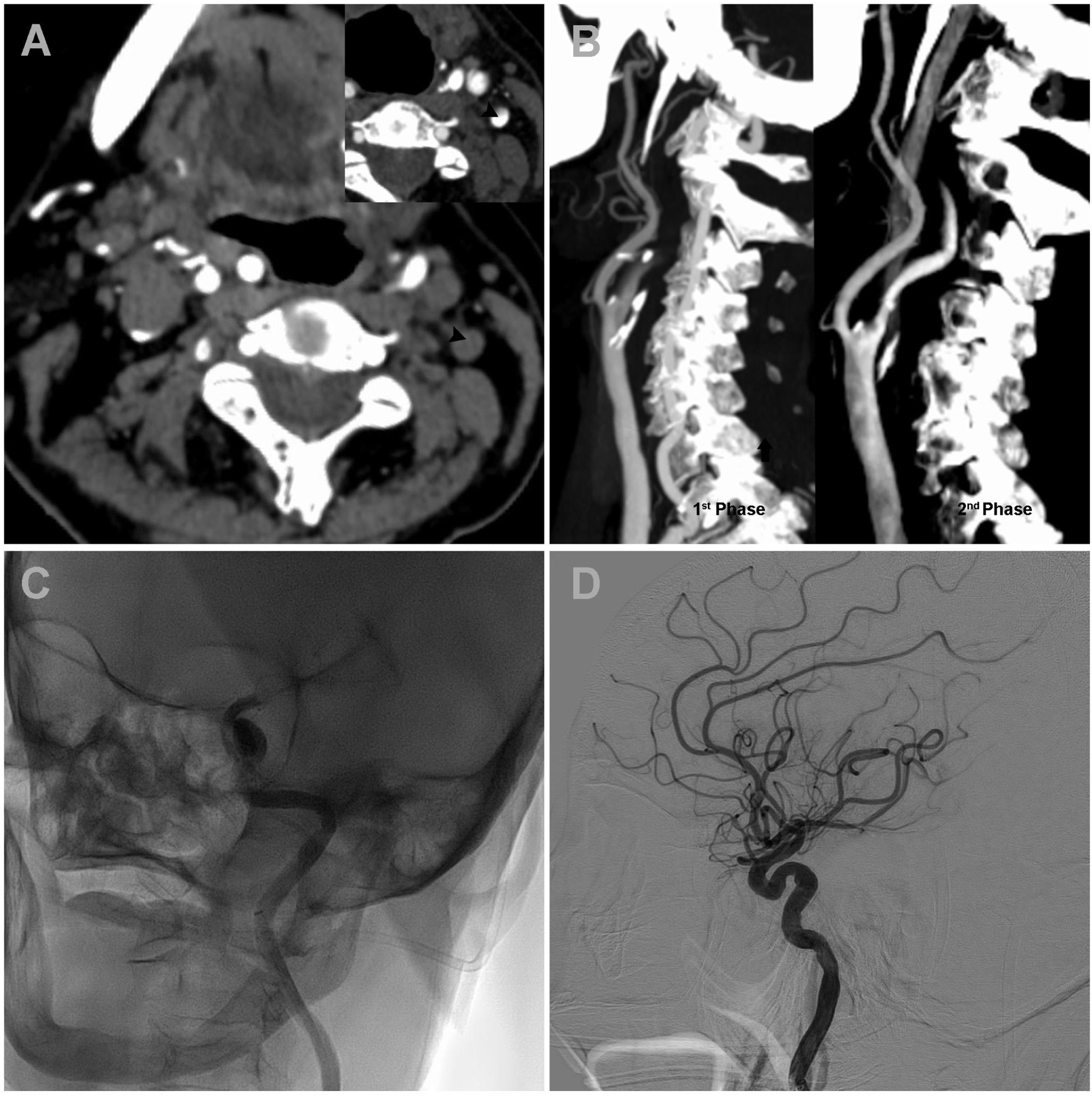
A representative case showing a patient of pseudo-occlusion at left ICA C1 segment with negative carotid ring sign but positive delayed contrast filling. Neither carotid ring sign nor contrast filling was detected on axial maximum intensity projection image in the 1^st^ phase of mCTA (arrow head, A), while delayed contrast filling was revealed in the 2^nd^ phase at left proximal C1 segment (arrow head, inset image in A). Sagittal images showed an apparent delayed ascending of contrast filling from proximal (1^st^ phase) to distal (2^nd^ phase) ICA C1 segment (B). DSA image shows a patent ICA at C3 segment and beyond (C). Successful recanalization with a modified Thrombolysis in Cerebral Infarction (eTICI) score of 2c was achieved after mechanical removal of thrombus at C5 segment (D).

**Table 2.** Comparison of diagnostic value of distinct CTA vessel features.

| | AUC (± SE) | Sensitivity<br>(95% CI) | Specificity<br>(95% CI) | p value (95% CI) | PPV (95% CI) | NPV (95% CI) | LR+ (95% CI) | LR- (95% CI) |
| --- | --- | --- | --- | --- | --- | --- | --- | --- |
| TO-Carotid ring sign | 0.86 (± 0.05) | 0.70 (0.60-0.79) | 0.94 (0.87-0.97) | 0.003 (0.59-0.85) | 0.90(0.80-0.95) | 0.80<br>(0.72-0.86) | 11.21<br>(5.4-23.1) | 0.31<br>(0.22-0.44) |
| PO-Delay contrast filling | 0.94 (± 0.04) | 0.95<br>(0.92-0.99 ) | 0.92 (0.73-0.99) | 0.000 (0.86-1.00) | 0.95(0.81-0.99) | 0.92(0.73-0.99) | 12.33<br>(3.25-46.80) | 0.06<br>(0.01-0.22) |

AUC indicate area under the curve; SE, standard error; CI, confidence region; PPV, positive predictive value; NPV, negative predictive value; LR, likelihood Ratios

## Discussion

The main finding of the study is that the presence of carotid ring sign from single-phase CTA or arterial phasic mCTA is a reliable marker to identify TO at ICA C1 segment. In addition, we confirmed the delayed contrast filling sign is a valuable marker to identify PO at the same site.

Carotid sign was defined as ring-like enhancement in carotid wall and/or intraluminal hypodensity on axial CTA sections. Firstly presented by Patrik Michel et al. that presence of carotid ring sign can differentiate acute symptomatic carotid occlusion from chronic asymptomatic occlusion^10^, they also proposed a pathophysiological explanation that the occurrence of the ring sign may be due to the contrast enhancement of the vasa vasorum in the viable arterial wall, and the luminal hypodensity corresponding to the absence of contrast uptake by the inherently vascular occluding thrombus^10^. The proposed pathophysiology helps to explain the high PPV of carotid ring sign in predicting carotid TO in this study. The main functions of the vasa vasorum are to deliver nutrients to the vessel wall and to remove waste products or noxious substances^11^. Adventitial vasa vasorum involves in the process of atherogenesis and atherosclerotic plaque progression^11^, and vasa vasorum enhancement recognized on CTA was strongly associated with symptomatic patients with carotid artery stenosis^12^.

Previous studies showed that 60%-70% causes of TO was atherosclerotic stenosis, 20%-30% was dissection and the remainder was carotid webs and cardiac emboli^13,14^. Our study showed 92% of cause of TO was atherosclerotic stenosis, 6% was dissection, 2% was cardioembolic. The difference could result from study samples and ethnics. Meanwhile, our study showed three quarters of TO had carotid ring sign, while very few (6%, 7 cases) of PO showed carotid ring sign. These 7 cases shared common atherosclerotic characteristics such as older age, hypertension and diabetes. Since most of these patients (87.5%, 6/7) also exhibited arterial fibrillation, we suggest TO should be considered with caution in elder patients with arterial fibrillation.

Delayed contrast filling on mCTA was reported to be a potent image marker in the identification of carotid occlusion site. As a contrast, single-phase CTA showed limited diagnostic value for the differentiation even for specialized radiologists^1^. Wareham, J. et al. first proposed to use multiphasic CTA to differentiated PO from TO, although the study failed to test the hypothesis^15^, but the study conducted by Choi, J.H. et al. did^8^. Delay time is one of the issues to account for the discrepancy. More than 40-second delay was employed in the former study, while there was only less than a 15-second delay in the latter. Extended delay time may cause the contrast to be washed out from target artery and failed to be detected by mCTA. Optimal CTA parameters to reveal hemodynamic abnormality for ICA pseudo-occlusion depend on different CT scanners. Consistent with the previous study, delayed contrast filling had high sensitivity and specificity and accuracy in differentiating PO from TO^8^. Of note, false positives of delayed contrast filling presented in TO patients may be due to, a) initial C1 high degree/near-occlusion stenosis; b) the culprit segment was located at or beyond middle segment of C1.

Carotid ring sign and delayed contrast filling each has its advantages and disadvantages. Carotid ring sign can be readily obtained from arterial phasic CTA without considering delay time, but its visual evaluation depends on raters’ experience and appropriate window level and window width. In comparison, although delay contrast filling from mCTA can easily spot ICA PO with a high sensitivity, the imaging quality may be affected by cardiac function, venous vasculature, motion artifacts and delay time.

Our study has limitations. First, the retrospective nature of this study may introduce uncontrolled bias. Second, all of the 200 patients with single-phase/arterial phasic CTA were included for the analysis of carotid ring sign, while only 69 patients with mCTA were analyzed for delayed contrast filling. The two datasets may be subject to uncontrolled/unadjusted variables. Third, since the carotid ring sign is visually identified in the study, it will be inevitably subject to the configuration of CT scanners, imaging acquisition process and inter-observer variance. Future studies are warranted to verify the generalizability of the findings.

In conclusion, we demonstrated that the carotid ring sign on sCTA is a reliable imaging marker to diagnose TO at proximal C1 ICA, and also confirmed the diagnostic value of delayed contrast filling sign on mCTA in diagnosing ICA PO.

## Data Availability

The data that support the findings of this study are available on request from the corresponding author.

